# Emergence of SARS-CoV-2 stains harbouring the signature mutations of both A2a and A3 clade

**DOI:** 10.1101/2021.02.04.21251117

**Authors:** Rakesh Sarkar, Anindita Banerjee, Shanta Dutta, Mamta Chawla-Sarkar

## Abstract

SARS-CoV-2 strains with both high transmissibility and potential to cause asymptomatic infection is expected to gain selective advantage over other circulating strains having either high transmissibility or ability to trigger asymptomatic infection. The D614G mutation in spike glycoprotein, the characteristic mutation A2a clade, has been associated with high transmissibility, whereas the A3 clade specific mutation L37F in NSP6 protein has been linked with asymptomatic infection. In this study, we performed a comprehensive mutational analysis of 3,77,129 SARS-CoV-2 genomes collected during January, 2020 to December, 2020 from all across the world for the presence of D614G and L37F mutations. Out of 3,77,129 SARS-CoV-2 strains analysed, 14, 598 (3.87%) were found to harbour both the D614G and L37F mutations. Majority of these double mutant SARS-CoV-2 strains were identified in Europe (11097) followed by North America (1915), Asia (980), Oceania (242), Africa (219), and South America (145). Geographical root surveillance revealed their first emergence during February-March in all the six continents. Temporal prevalence analysis from February, 2020 to December, 2020 showed a gradual upsurge in their frequencies worldwide, which strongly demonstrated the adaptive selection of these double mutants. Evolutionary analysis depicted that these double mutants emerged as a new clade in the dendrogram (named as A2a/3), and were sub-divided into four distinct clusters (Cluster I, II, III and IV) according to different sets of coexisting mutations. The frequency distribution pattern showed the global predominance of cluster III (41.42%), followed by cluster IV (23.31%), cluster II (21.02%) and cluster I (14.25%). Overall, our study highlighted the emergence of a unique phylogenetic clade encompassing the double-mutant SARS-CoV-2 strains which may provide a fitness advantage during course of virus evolution.

## 1. Introduction

The ongoing global pandemic of coronavirus disease 2019 (COVID-19) caused by severe acute respiratory syndrome coronavirus 2 (SARS-CoV-2) was first reported from Wuhan, China in late December 2019. As of now (29^th^ January, 2021), the virus had spread to 221 countries around the world, infecting around 102,041,120 people with 2,201,044 deaths (https://www.worldometers.info/coronavirus/). This rapid surge of infection within one year of inception has overwhelmed existing health care systems with patients and imposed a significant hostile effect on social and economic activities worldwide.

The major challenges in controlling the rapidly mutating SARS-CoV-2 virus is its high transmissibility and low virulence. Some of the mutations which accumulated during the early pandemic-phase enabled virologists to classify the circulating SARS-CoV-2 strains into different lineages and clades (Gomez-Carballa et al., 2020). Among these, the D614G mutation emerged in the spike glycoprotein, the key protein involved in the binding of SARS-CoV-2 to host cell receptor angiotensin-converting enzyme-2 (ACE-2). D614G is the characteristic mutation of A2 clade which was first collected during January, 2020 from Germany. Gradually, the D614G mutants were seen to predominate in rest of Asia, Europe, North America and Oceania (Gomez-Carballa et al., 2020). This rapid spreading led scientists to address whether the D614G mutation could render SARS-CoV-2 more infectious (Korber et al., 2020; Plante et al., 2020; Omotuyi et al., 2020; Jiang et al., 2020; Zhang et al., 2020; Gobeil et al., 2020). Recent studies provided both experimental and clinical evidence in favour of high infectivity of SARS-CoV-2 isolates harbouring the D614G mutation compared to the prototype strain, however no co-relation with disease severity was observed (Korber et al., 2020, Plante et al., 2020; Zhang et al., 2020). It has been confirmed that G614 variant grow to higher titers in human lung cell lines due to enhanced infectivity (Korber et al., 2020, Plante et al., 2020; Zhang et al., 2020). Patients infected with D614G mutants were found to have higher loads of viral RNA compared to patients infected with the prototype strain (Korber et al., 2020). In addition, D614G substitution has been shown to enhance the furin cleavage efficiency at S1/S2 junction of S glycoprotein (Gobeil et al., 2020) which is important for SARS-CoV-2 infection (Bestle et al., 2020; Shang et al., 2020).

Another mutation that emerged in China during January, 2020 is L37F within the NSP6 protein, the characteristic mutation of the A3 clade. Very soon, this mutant emerged in the United States followed by Italy and France. Though L37F variants had arrived in USA and UK at very early phase of the pandemic, they could not over-rule the other preponderant lineages like A2a due their poor transmissibility (Aiewsakun et al., 2020). Two recent studies have shown that SARS-CoV-2 strains isolated from asymptomatic COVID patients had L37F mutation in the NSP6 protein, suggesting a strong correlation between L37F mutation and weakened SARS-CoV-2 virulence (Wang et al., 2020; Aiewsakun et al., 2020). By using variety of simulation and network models it has also been demonstrated that L37F mutation disrupts the folding stability of NSP6 which may have compromised the virus’s ability to undermine the innate cellular defense against viral infection via autophagy regulation (Wang et al., 2020).

Strains harbouring these D614G and L37F mutations are expected to have high transmissibility and weak virulence thus causing asymptomatic infection, which might allow them a selective advantage and to spread very rapidly in the population. With this hypothesis, in the present study, we analysed the spatial and temporal distribution of strains having both D614G and L37F mutations and also evaluated their phylogenetic characteristics.

## 2. Material and Methods

In total, we downloaded 3,77,129 genome sequences of SARS-CoV-2 strains from the global initiative of sharing all influenza data (GISAID) (https://www.gisaid.org/). All the viral samples collected till 31^st^ December, 2020 and corresponding genome sequences submitted on or before 26^th^ January, 2020 were included. 10 clade-specific reference gene sequences of SARS-CoV-2 were also downloaded from GISAID for dendrogram construction and subsequent analysis of lineage distribution and clade formation. The reference genome sequence of SARS-CoV-2 isolate Wuhan-Hu-1 (MN908947.3) used for mutational analysis was downloaded from National Center for Biotechnology Information (NCBI). For phylogenetic analysis multiple genome sequence alignment was done with online alignment program MAFT version 7 (https://mafft.cbrc.jp/alignment/server/). The whole genome phylogenetic dendrogram was constructed using the MAFT alignment file by MEGA, version X (Molecular Evolutionary Genetics Analysis), employing the maximum-likelihood statistical method (at 1000 bootstrap replicates) and using the best fit nucleotide substitution model. To identify the amino acid change, genome sequences were translated using TRANSEQ (Transeq Nucleotide to Protein Sequence Conversion Tool, EMBL-EBI, Cambridgeshire, UK).

## 3. Results

### 3.1. Gradual upsurge of SARS-CoV-2 strains harbouring D614G and L36F mutations with growing months across the world

During mutational and phylogenetic analysis of circulating clades in India (Sarkar et al., 2020), we noticed SARS-CoV-2 strains harbouring both D614G and L37F mutations. Based on this observation, we further performed mutational analysis of 377,129 SARS-CoV-2 genome sequences, deposited in GISAID from countries of six different continents around the world, for the presence of both D614G and L37F mutations. Most of the genome sequences analyzed were submitted from Europe (n=246,448) followed by North America (n=84,448), Asia (n=17,771), Oceania (n=17,531), Africa (n=5,804) and South America (n=5,127) (**Supplimentary Figure 1A**). Genomes of 14,598 SARS-CoV-2 strains among 377,129 (3.87%) were found to harbour both D614G and L37F mutations (**Figure 1A**). Maximum number of the double mutant SARS-CoV-2 strains were detected in Europe (11097) followed by North America (1915), Asia (980), Oceania (242), Africa (219), and South America (145). (**Supplimentary Figure 1B**). However, continent wise percentage positivity analysis showed the highest predominance of these double mutant SARS-CoV-2 strains in Asia (980/17771, 5.51%) followed by Europe (11097/246448, 4.5%), Africa (219/5804, 3.77%), South America (145/5127, 2.82%), North America (1915/84448, 2.26%) and Oceania (242/17531, 1.38%) (**Figure 1B**). Furthermore, we analysed the temporal percentage positivity of the double mutant SARS-CoV-2strain among all the strains collected worldwide from the time they first appeared in February, 2020 to December, 2020. Surprisingly, we observed a steady rise in the frequency of the double mutant SARS-CoV-2 strain from February (0.14%) to October (7.65%) followed by decline in November (5.14%) and December (3.01%) worldwide (**Figure 1C**). However, the trend line showed the gradual increase in the frequency of double mutant over time. Decreased percentage of the double mutant strain in November and December was due to low number of sequence availability in GISAID. The actual percentage positivity may be obtained after 3 to 4 months when all the samples collected during November and December will be sequenced. Analysis of temporal prevalence among different continents also revealed an increasing trend in the frequency of double mutant strains from February to December (**Figure 1D-1I**). Overall, our results demonstrated a gradual upsurge of SARS-CoV-2 strains having both D614G and L37F mutations with time.

**Figure 1:**
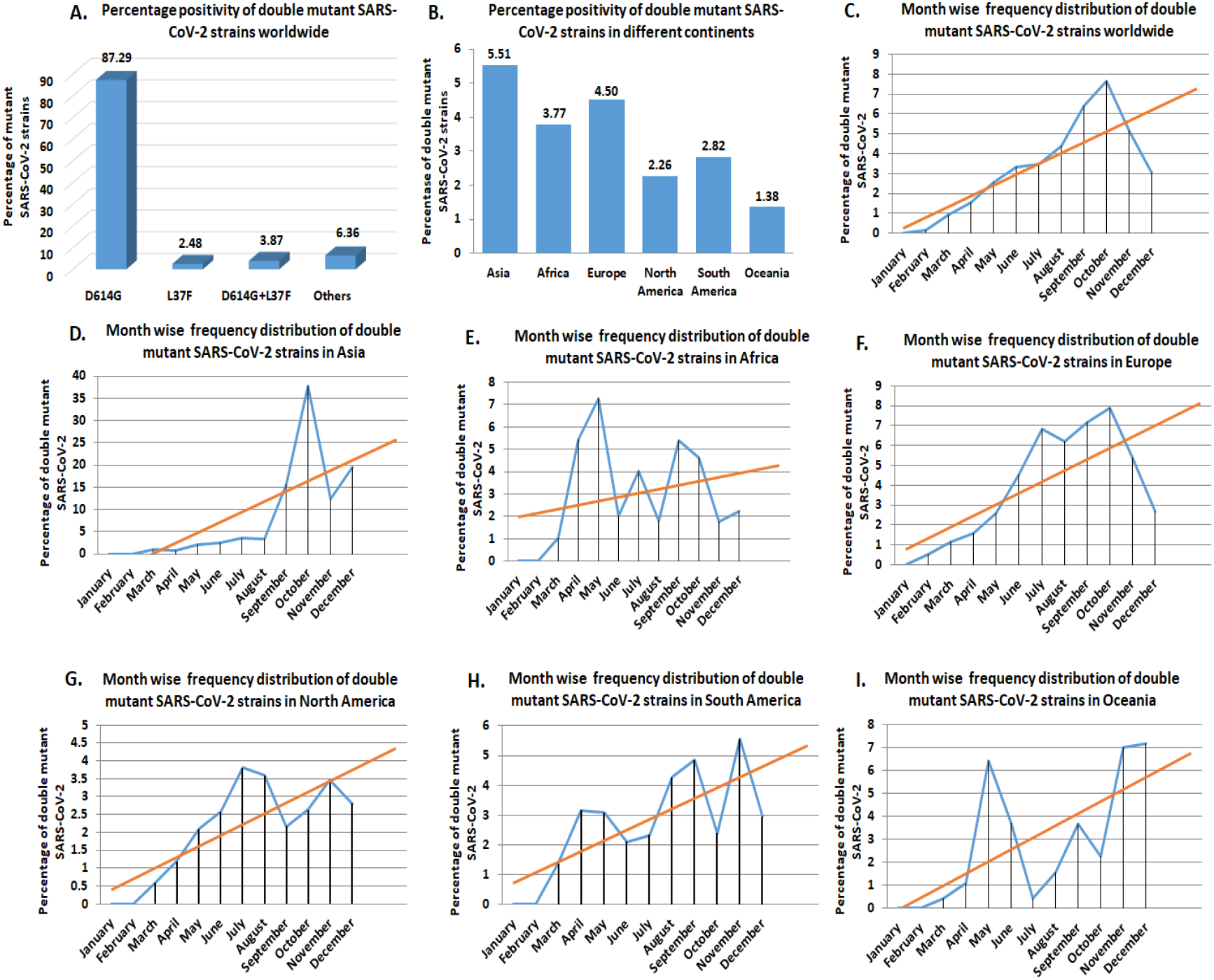
Spatial and temporal distribution of SARS-CoV-2 strains harbouring both D614G and L37F mutations. (A) The bar graph representing the percentage positivity of the SARS-CoV-2 strains having D614G or L37F or both mutations worldwide. (B) The bar graph representing the percentage positivity of the double mutant SARS-CoV-2 strains among different continents. (C) Graph representing the temporal frequencies of the double mutant SARS-CoV-2 strains worldwide. (D-I) Temporal frequency analysis of the double mutant SARS-CoV-2 strains in Asia (D), Africa (E), Europe (F), North America (G), South America (H) and Oceania (I).

### 3.2. Phylogenetic analysis of the double mutant SARS-CoV-2 strains revealed four different clusters

Next, we performed the phylogenetic analysis of these double mutant strains. Phylogenetic dendrogram was constructed on the basis of whole genome sequences of 107 strains consisting of 11 clade-specific strains and 96 randomly selected double-mutant strains from 14598 strains encompassing all the continents. The dendrogram revealed that these 96 double-mutants clustered among themselves, revealing the emergence of a unique clade (named here as Clade A2a/3) which was distant from all the 11 other clade specific strains. The new clade A2a/3 strains exhibited 4 distinct clusters within themselves in the dendrogram (cluster I marked in pink colour-21 strains, cluster II marked in blue colour-29 strains, cluster-III marked in green colour-27 strains and cluster IV marked in red colour-19 strains). This newly emerging clade A2a/3 had maximum genetic closeness to the A2a clade specific strains (97-99% DNA homology) compared to the other clade strains in the tree. The B2 clade was genetically farthest to this A2a/3 clade (**Figure 2**).

**Figure 2:**
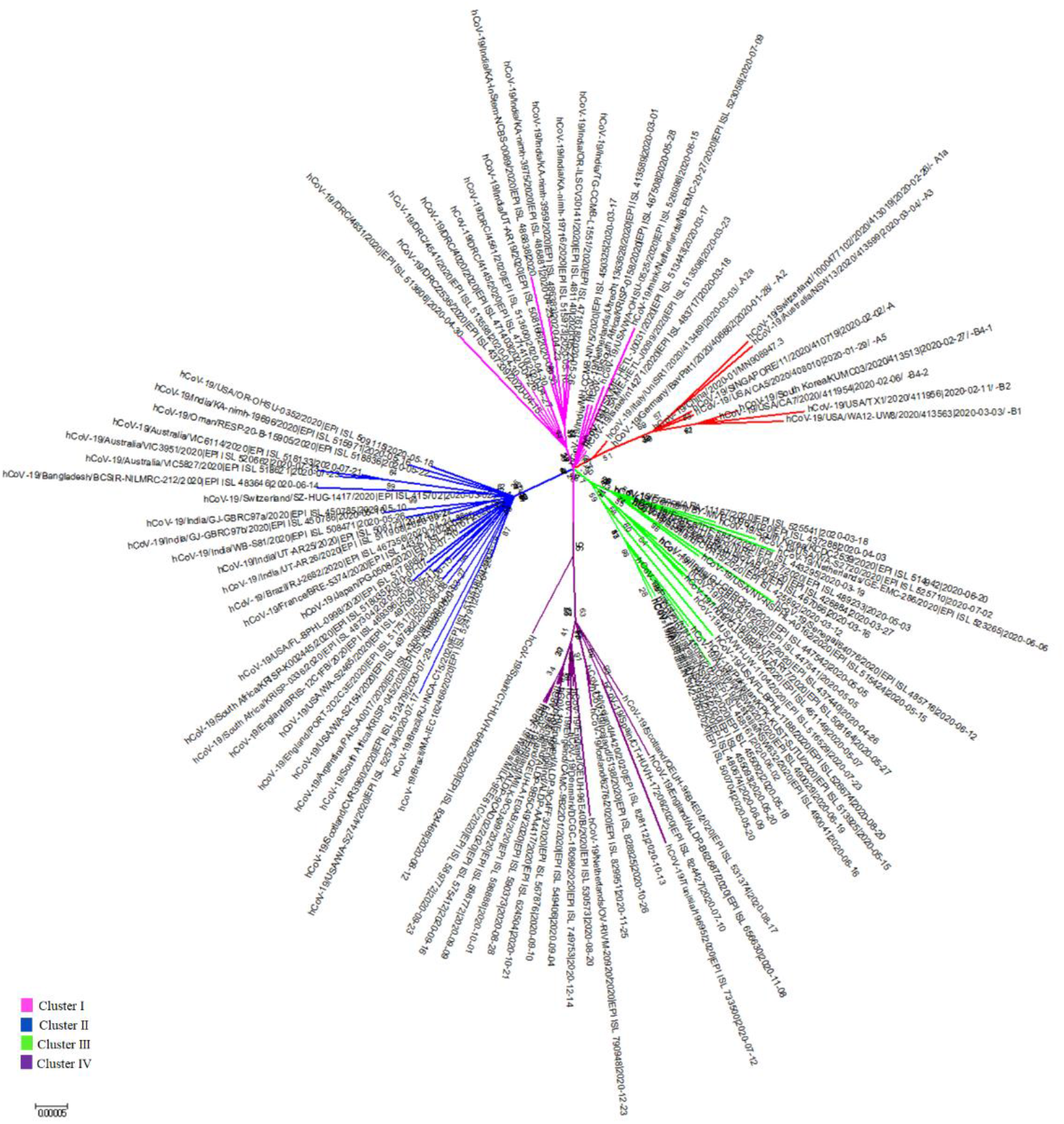
Molecular phylogenetic analysis by maximum likelihood method. Phylogenetic dendrogram based on whole genome sequences of 96 representative strains along with 10 clade specific known strains and the prototype O-clade strain (MN908947). Scale bar was set at 0.00005 nucleotide substitution per site. The best fit model used for constructing the phylogenetic dendrogram was General Time Reversible Model (GTR).

### 3.3. Mutational analysis of the four clusters and their frequency distribution among different continents

We performed the whole genome mutational analysis of 96 double mutant strains encompassing the four different clusters with compared to the reference genome MN908947.3 to uncover the unique mutations that separate them into distinct clusters. Mutational analysis revealed that Cluster I strains harboured five common mutations; 241C>T in 5’-UTR, 3037C>T (F106F) in NSP3, 11083G>C (L37F) in NSP6, 14408C>T (P323L) in RdRp and 23403 (D614G) in S glycoprotein. Strains of Cluster II, Cluster III and Cluster IV were found to contain additional 25563G>T (Q57H) in ORF3a, 28881GGG>CCA (RG203KR) in N protein and 22227C>T (A222V) in S glycoprotein respectively (**Table 1**). Furthermore, whole genome mutational analysis of the remaining 14502 double mutant strains was done to check for the presence of the aforementioned cluster specific mutations. The frequency distribution pattern of the 4 different clusters showed the maximum occurrence of cluster III (6047/14598, 41.42%), followed by cluster IV (3403/14598, 23.31%), cluster II (3068/14598, 21.02%) and cluster I (2080/14598, 14.25%) across the world (**Figure 3A**). The 2080 strains belonging to cluster I were maximally found across Europe (1513/2080, 72.74%), followed by North America (366/2080, 17.6%), Africa (107/2080, 5.14%), Asia (50/2080, 2.4%), Oceania (29/2080, 1.4%) and South America (15/2080, 0.72%). The frequency of cluster II strains peaked across Europe (1444/3068, 47.06%) followed by North America (1181/3068, 38.5%), Asia (346/3068, 11.27%), Oceania (65/3068, 2.11%), South America (20/3068, 0.65%) and Africa (12/3068, 0.4%). The strains of cluster III were found to predominate in Europe (4748/6047, 78.51%), followed by Asia (584/6047, 9.65%), North America (368/6047, 6.08%), Oceania (138/6047, 2.28%), South America (110/6047, 1.81%) and Africa (99/6047, 1.63%). The frequency of cluster IV strains peaked in Europe (3392/3403, 99.67%), followed by Oceania (10/3403, 0.3%) and Africa (1/3403, 0.03%). Strains of cluster IV were not found in North America, South America and Asia (**Figure 3B**). In summary, all the clusters strains maximally predominated in Europe.

**Table 1:** Co-existing mutations present within the four different clusters of the double mutant SARS-CoV-2 strains.

| Cluster | Co-existing mutations |  |  |  |  |  |  |  |
| --- | --- | --- | --- | --- | --- | --- | --- | --- |
|  | 241<br>C>T<br>5'-UTR | 23403 A>G<br>D614G<br>S Glycoprotein | 14408 C>T<br>P323L<br>RdRp/NSP12 | 3037<br>C>T<br>F106F<br>NSP3 | 11083<br>G>C<br>L37F<br>NSP6 | 25563<br>G>T<br>Q57H<br>ORF3a | 28881 GGG>AAC<br>RG203KR<br>Nucleoprotein | 22227C>T<br>A222V<br>S Glycoprotein |
| Cluster I | ✓ | ✓ | ✓ | ✓ | ✓ |  |  |  |
| Cluster II | ✓ | ✓ | ✓ | ✓ | ✓ | ✓ |  |  |
| Cluster III | ✓ | ✓ | ✓ | ✓ | ✓ |  | ✓ |  |
| Cluster IV | ✓ | ✓ | ✓ | ✓ | ✓ |  |  | ✓ |

**Figure 3:**
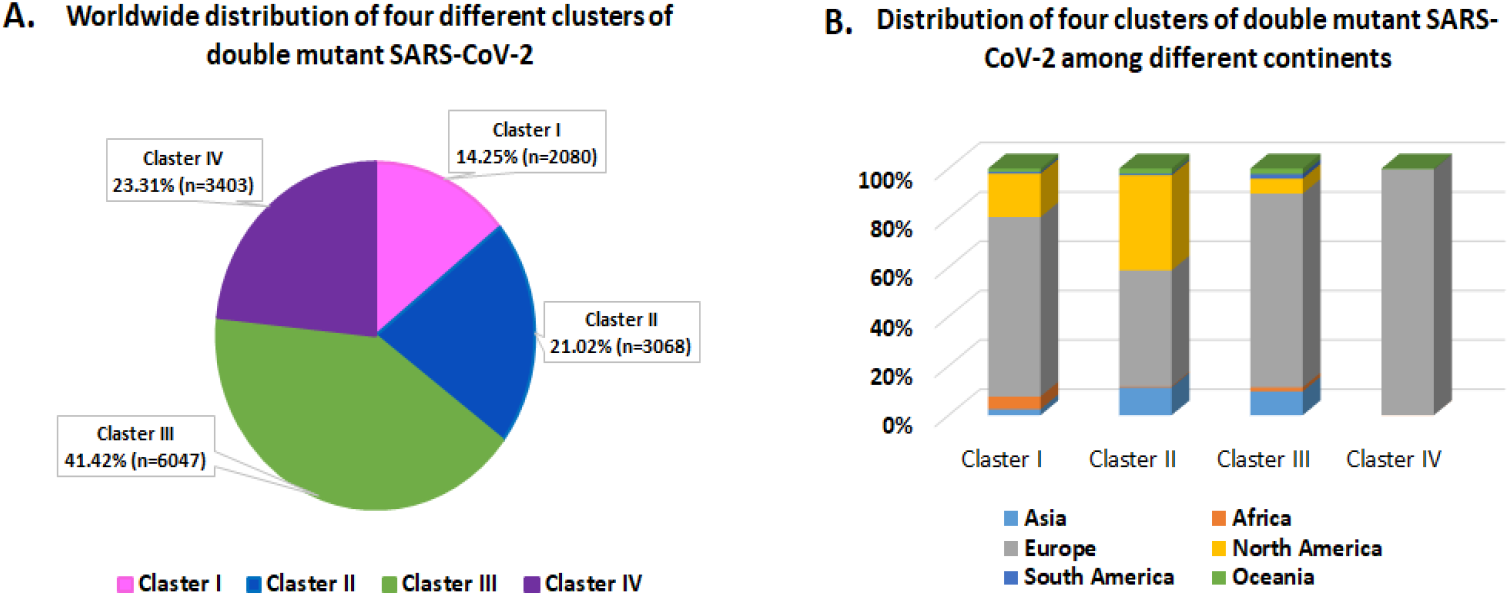
Frequency distribution of four distinct clusters of the double mutant SARS-CoV-2 strains. (A) The pie chart representing the global frequency distribution of the four distinct clusters of the double mutant SARS-CoV-2 strains. (B) A visualization of frequency distribution of the four distinct clusters of the double mutant SARS-CoV-2 strains among different continents.

### 3.4. Geographical root monitoring of the four clusters of emerging A2a/3 clade

In-depth analyses of the geographical origin of the A2a/3 clade revealed that all the four cluster strains were first collected and reported from Europe. Cluster I originated foremost in Europe (Italy on February 24), followed by North America (USA on March 10, 2020), Asia (Hong Kong on March 14, 2020), Oceania (Australia on March 24, 2020), South America (Chile on March 25, 2020), and Africa (Morocco on March 31, 2020). Cluster II originated first in Europe (France on March 10, 2020), followed by North America (USA on March 13, 2020), Asia (Singapore on March 15, 2020), Oceania (Australia on March 18, 2020), Africa (Senegal on March 20, 2020 and South America (Trinidad on August 14, 2020). Strains of cluster III appeared first within Europe (Ireland on February 20, 2020), followed by Asia (Israel on February 27, 2020), South America (Brazil on March 4, 2020), Oceania (Australia on March 23, 2020), Africa (South Africa on March 27, 2020) and North America (Canada on March 28, 2020). Strains of the cluster IV was seen to emerge first in Europe (Spain on June 3, 2020)), followed by Africa (Tunicia on July 12, 2020) and Oceania (Australia on August 20) (**Table 2**).

**Table 2:** Geographical root monitoring of the four different clusters of double mutant SARS-CoV-2 strains.

| Continents | Cluster-I |  |  | Cluster- II |  |  | Cluster -III |  |  | Cluster- IV |  |  |
| --- | --- | --- | --- | --- | --- | --- | --- | --- | --- | --- | --- | --- |
|  | Collecti on date | Count ry | Accession ID | Collecti on date | Countr y | Accession ID | Collecti on date | Count ry | Accession ID | Collecti on date | Count ry | Accession ID |
| Asia | 14.03.2020 | Hong Kong | EPI_ISL_430063 | 15.03.2020 | Singapore | EPI_ISL_548989 | 27.02.2020 | Israel | EPI_ISL_649094 | Not found till 31<sup>st</sup> December, 2020 |  |  |
| Africa | 31.03.2020 | Morocco | EPI_ISL_728221 | 20.03.2020 | Senegal | EPI_ISL_420079 | 27.03.2020 | South Africa | EPI_ISL_436686 | 12.07.2020 | Tunisia | EPI_ISL_733500 |
| Europe | 24.02.2020 | Italy | EPI_ISL_856884 | 10.03.2020 | France | EPI_ISL_569250 | 20.02.2020 | Ireland | EPI_ISL_605797 | 23.06.2020 | Spain | EPI_ISL_537739 |
| North America | 10.03.2020 | USA | EPI_ISL_632047 | 13.03.2020 | USA | EPI_ISL_424174 | 28.03.2020 | Canada | EPI_ISL_538330 | Not found till 31<sup>st</sup> December, 2020 |  |  |
| South America | 25.03.2020 | Chile | EPI_ISL_801654 | 14.08.2020 | Trinidad | EPI_ISL_717696 | 04.03.2020 | Brazil | EPI_ISL_416034 | Not found till 31<sup>st</sup> December, 2020 |  |  |
| Oceania | 24.03.2020 | Australia | EPI_ISL_451602 | 18.03.2020 | Australia | EPI_ISL_639795 | 23.03.2020 | Australia | EPI_ISL_794688 | 20.08.2020 | Australia | EPI_ISL_593721 |

## 4. Discussion

High transmissibility and triggering of elusive asymptomatic infections are Achilles heel to the current evasion strategies to control SARS-CoV-2 infection, which encompasses symptom-based testing, contact tracing, isolation and quarantine measures. Virus transmission rate and degree of disease severity could be associated with various host as well as viral factors. Host factors include age, CD4+ and CD8+ T cell counts and level of IL-6 and IL-8 secretion, whereas viral factors include genomic diversity which could generate more pathogenic and virulent strains (Guan et al., 2020; Yang et al., 2020; Zhang et al., 2020). Several recent studies have focused on potential genetic variations across the SARS-CoV-2 genome that could be linked to high transmissibility and occurrence of asymptomatic infections. Two single nucleotide polymorphisms (SNPs) 23403A>G and 11083G>T were claimed to be associated high transmissibility and asymptomatic infection, respectively (Korber et al., 2020; Plante et al., 2020; Wang et al., 2020; Aiewsakun et al., 2020).

Spike gene is the basis of host-immune selective pressure, and antibodies produced against it inhibit viral attachment and entry into the host-cells (Korber et al., 2020). The SNP 23403A>G is responsible for the D614G mutation in the spike glycoprotein (614^th^ Aspartic acid changes to Glycine) that plays an essential role in binding to the host ACE2 receptors determining host tropism (Fung et al., 2019). This mutation was seen to first emerge within Germany during January, 2020 and eventually outcompeted the other pre-existing strains worldwide, including the ancestral sub-type. Soon it was seen to get established as the cardinal strain of the global pandemic, underscoring that this mutation confers efficient human to human transmission ability (Grubaugh et al., 2020). Among all the spike variants, although D614G mutant enhances viral transmissibility, nevertheless it maintains neutralization vulnerability by antisera against the wild-type virus (Weissman et.al., 2020). Reports show that the mutant variety is comparatively stable than the wild type, which was circulating at low frequency in March, but quickly upsurged through April and May, revealing a transmission advantage over other co-circulating strains (Zhang et.al., 2020). Homology modeling revealed that this D614G mutation might gain a competitive advantage at the furin binding domain which was a probably responsible for the upsurge (Tang et.al., 2020). However, the clinical manifestations of this mutant strain is yet to be completely annotated (Eaaswarkhanth et al., 2020; Korber et al., 2020).

It has been reported that D614G mutation accelerates SARS-CoV-2 replication in lung epithelial cells and primary airway cells (Plante et.al., 2020). It also creates an additional serine protease (Elastase) cleavage site near the S1-S2 spike junction that may enhance virus membrane fusion by several fold (Bhattacharya et.al., 2020). An interesting question arises that why do these mutant viruses reveal more transmissibility without causing any disease severity? It is possible that other viral/ host factors might diminish the rate and efficiency of intra-host replication of these mutant strains. In silico study revealed that it is most probably neutral to the protein function (Isabel et.al., 2020). Reports also claim that in human lung epithelial cell-lines, the D614G mutation increased transduction and resistance to cleavage (Daniloski et.al., 2020). Hence, the efficacy of the developing vaccines in protecting against the emerging mutant strain remains questionable.

The L37F mutation in the non-structural protein 6 (NSP6) corresponding to the ‘A3’ phylogenetic clade was associated maximally with cases from UK (Forster et al., 2020). The first confirmed COVID-19 positive case was detected in Singapore on January 23, 2020, and thereafter within one week, the first strain with L37F mutation was reported from Singapore on January 29,2020. Initially across Singapore, a huge proportion of the L37F mutants was observed, and the prevalence of this mutation might be associated with the very low death ratio in Singapore. Analyses of various sequences with asymptomatic and symptomatic patient history, and was able to relate this mutation to the asymptomatic patients. By tracking the geographical distribution, this mutation could be correlated with the death ratio in countries like Singapore, United States, United Kingdom, etc, strongly concluding the association between the asymptomatic infection and viral hypotoxicity. Study on the worldwide-dynamics of L37F mutation focused on its decaying tendency, indicating that it hinders viral transmissibility. Furthermore, the analysis of the distribution of L37F mutants in different age-groups and genders shows that this mutation does not depict a host-dependent behaviour and is independent of age as well as gender. L37F destabilizes the structure of NSP6 protein rendering it energetically less stable. The rigidity and flexibility index along with many network modelling analyses revealed that L37F might have compromised the NSP6 function, leading to a relatively weaker coronavirus subtype. As one of the most conservative proteins of SARS-CoV-2, NSP6 plays a crucial role in regulation of viral autophagy. Therefore, this destabilized mutation can compromise the efficiency of NSP6 to help in viral protein folding, assembly of virions, and virus replication cycle, which underscores its relevance among the asymptomatic infections (Wang et.al., 2020).

Our analyses of sequences submitted from different continents revealed the maximum prevalence of the double-mutant strains across Europe. The significant temporal increase in the double mutant frequency within 10 months clearly demonstrated their adaptive selection across the world as well as in different continents (Figure 1). Phylogenetic analyses revealed that the double-mutant strains emerged as a unique clade in the phylogenetic dendrogram (named as A2a/3 clade) which had maximum identity to the ancestral A2a clade (Figure 2). This emerging clade was again sub-divided into 4 clusters within themselves, each having their specific sets of mutation (Table 1) and geographical origin across the globe (Table 2). Mutational analysis revealed that the emergence of L37F mutation within the G clade, GH clade, GR clade and GV clade (Mercatelli et al., 2020) might give rise to the evolution of cluster I, cluster II, cluster III and cluster IV respectively. We did not find the emergence of D614G or L37F mutation within the new UK variant (Rambaut et al., 2020).

During the rapid surge of COVID-19 pandemic, understanding the evolution of these double-mutant SARS-CoV-2 viruses is extremely crucial for the disease control and prevention. To spread, a virus must multiply within the host to ensure transmission, while simultaneously avoiding host morbidity or death. Therefore, during virus evolution the transmissibility is usually increased whereas the pathogenicity becomes reduced. Constant monitoring of co-existence of both these mutations will be pivotal in tracking the movement of the virus between individuals and across different geographical areas, and will allow for scalable phylogenetic analyses and real time tracking of specific mutations. Worldwide dispersal and increasing frequency of theses double-mutant strains indicates a strong selective/ fitness advantage, but might also arise on account of a random founder effect. The significant differences in the growth and size of phylogenetic clusters of these double-mutants indicate a need for continuous monitoring of their frequency. Moreover, the mutational impact on viral spread and vaccine efficacy needs to be studied in depth.

## Data Availability

Available on request

## Acknowledgement

The authors acknowledge the intramural financial support by ICMR, India. RS is supported by Senior Research Fellowship from University Grants Commission (UGC), India.

## Figure legend

**Supplimentary Figure 1:**
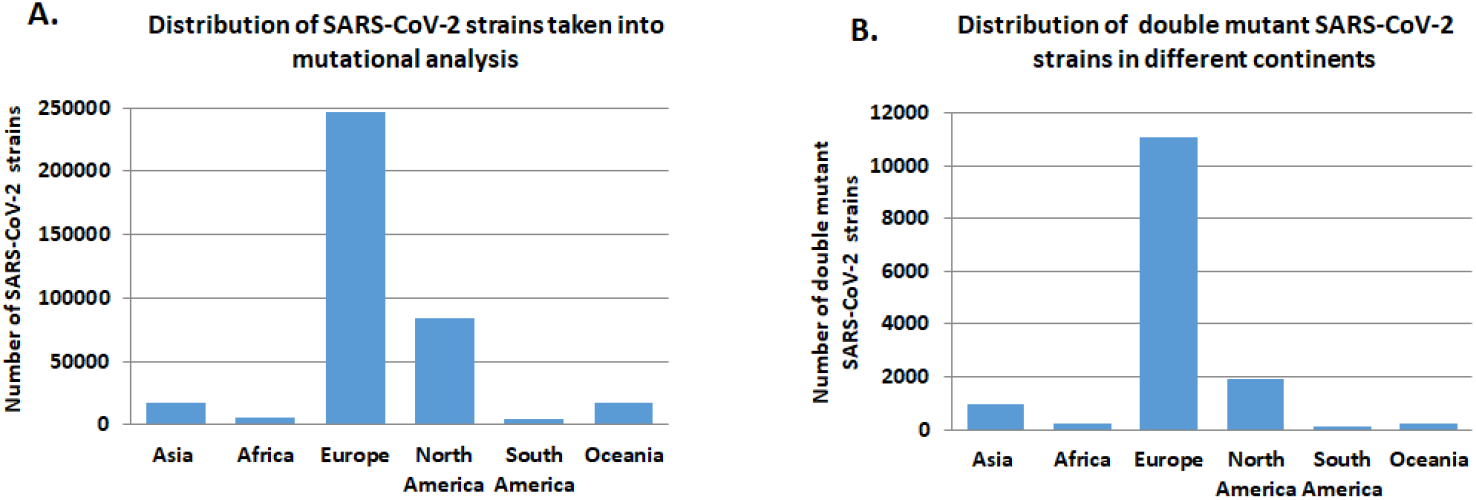
(A) Graph representing the frequency distribution of SARS-CoV-2 strains taken into mutational analysis among different continents. (B) Graph demonstrating the frequency distribution of SARS-CoV-2 strains harbouring both D614G and L37F mutations in different continents.

## References

Aiewsakun, P., Wongtrakoongate, P., Thawornwattana, Y., Hongeng, S. and Thitithanyanont, A., 2020. SARS-CoV-2 genetic variations associated with COVID-19 severity. medRxiv.

Bestle, D., Heindl, M.R., Limburg, H., Pilgram, O., Moulton, H., Stein, D.A., Hardes, K., Eickmann, M., Dolnik, O., Rohde, C. and Klenk, H.D., 2020. TMPRSS2 and furin are both essential for proteolytic activation of SARS-CoV-2 in human airway cells. Life science alliance, 3(9).

Bhattacharyya, C., Das, C., Ghosh, A., Singh, A.K., Mukherjee, S., Majumder, P.P., Basu, A. and Biswas, N.K., 2020. Global Spread of SARS-CoV-2 Subtype with Spike Protein Mutation D614G is Shaped by Human Genomic Variations that Regulate Expression of TMPRSS2 and MX1 Genes. bioRxiv.

Daniloski, Z., Guo, X. and Sanjana, N.E., 2020. The D614G mutation in SARS-CoV-2 Spike increases transduction of multiple human cell types. BioRxiv.

Eaaswarkhanth, M., Al Madhoun, A. and Al-Mulla, F., 2020. Could the D614 G substitution in the SARS-CoV-2 spike (S) protein be associated with higher COVID-19 mortality?. International Journal of Infectious Diseases.

Forster, P., Forster, L., Renfrew, C. and Forster, M., 2020. Phylogenetic network analysis of SARS-CoV-2 genomes. Proceedings of the National Academy of Sciences, 117(17), pp. 9241–9243.

Fung, T.S. and Liu, D.X., 2019. Human coronavirus: host-pathogen interaction. Annual review of microbiology, 73, pp. 529-557.

Gobeil, S.M., Janowska, K., McDowell, S., Mansouri, K., Parks, R., Manne, K., Stalls, V., Kopp, M.F., Henderson, R., Edwards, R.J. and Haynes, B.F., 2020. D614G mutation alters SARS-CoV-2 spike conformation and enhances protease cleavage at the S1/S2 junction. Cell reports, p.108630.

Gómez-Carballa, A., Bello, X., Pardo-Seco, J., Martinón-Torres, F. and Salas, A., 2020. Mapping genome variation of SARS-CoV-2 worldwide highlights the impact of COVID-19 super-spreaders. Genome Research, 30(10), pp. 1434–1448.

Grubaugh, N.D., Hanage, W.P. and Rasmussen, A.L., 2020. Making sense of mutation: what D614G means for the COVID-19 pandemic remains unclear. Cell, 182(4), pp. 794–795.

Guan, W.J., Ni, Z.Y., Hu, Y., Liang, W.H., Ou, C.Q., He, J.X., Liu, L., Shan, H., Lei, C.L., Hui, D.S. and Du, B., 2020. Clinical characteristics of coronavirus disease 2019 in China. New England journal of medicine, 382(18), pp. 1708–1720.

Isabel, S., Graña-Miraglia, L., Gutierrez, J.M., Bundalovic-Torma, C., Groves, H.E., Isabel, M.R., Eshaghi, A., Patel, S.N., Gubbay, J.B., Poutanen, T. and Guttman, D.S., 2020. Evolutionary and structural analyses of SARS-CoV-2 D614G spike protein mutation now documented worldwide. Scientific reports, 10(1), pp. 1–9.

Jiang, X., Zhang, Z., Wang, C., Ren, H., Gao, L., Peng, H., Niu, Z., Ren, H., Huang, H. and Sun, Q., 2020. Bimodular effects of D614G mutation on the spike glycoprotein of SARS-CoV-2 enhance protein processing, membrane fusion, and viral infectivity. Signal transduction and targeted therapy, 5(1), pp. 1–3.

Korber, B., Fischer, W.M., Gnanakaran, S., Yoon, H., Theiler, J., Abfalterer, W., Hengartner, N., Giorgi, E.E., Bhattacharya, T., Foley, B. and Hastie, K.M., 2020. Tracking changes in SARS-CoV-2 Spike: evidence that D614G increases infectivity of the COVID-19 virus. Cell, 182(4), pp. 812–827.

Mercatelli, D. and Giorgi, F.M., 2020. Geographic and genomic distribution of SARS-CoV-2 mutations. Frontiers in microbiology, 11, p. 1800.

Omotuyi, I.O., Nash, O., Ajiboye, O.B., Iwegbulam, C.G., Oyinloye, E.B., Oyedeji, O.A., Kashim, Z.A. and Okaiyeto, K., 2020. Atomistic simulation reveals structural mechanisms underlying D614G spike glycoprotein-enhanced fitness in SARS-COV-2. Journal of computational chemistry, 41(24), pp. 2158–2161.

Plante, J.A., Liu, Y., Liu, J., Xia, H., Johnson, B.A., Lokugamage, K.G., Zhang, X., Muruato, A.E., Zou, J., Fontes-Garfias, C.R. and Mirchandani, D., 2020. Spike mutation D614G alters SARS-CoV-2 fitness. Nature, pp. 1–6.

Rambaut, A., Loman, N., Pybus, O., Barclay, W., Barrett, J., Carabelli, A., Connor, T., Peacock, T., Robertso, D.L., Volz, E., 2020. Preliminary genomic characterisation of an emergent SARS-CoV-2 lineage in the UK defined by a novel set of spike mutations. https://virological.org/t/preliminary-genomic-characterisation-of-an-emergent-sars-cov-2-lineage-in-the-uk-defined-by-a-novel-set-of-spike-mutations/563.

Sarkar, R., Mitra, S., Chandra, P., Saha, P., Banerjee, A., Dutta, S. and Chawla-Sarkar, M., 2021. Comprehensive analysis of genomic diversity of SARS-CoV-2 in different geographic regions of India: An endeavour to classify Indian SARS-CoV-2 strains on the basis of co-existing mutations. Arch Virol.

Shang, J., Wan, Y., Luo, C., Ye, G., Geng, Q., Auerbach, A. and Li, F., 2020. Cell entry mechanisms of SARS-CoV-2. Proceedings of the National Academy of Sciences, 117(21), pp. 11727–11734.

Tang, L., Schulkins, A., Chen, C.N., Deshayes, K. and Kenney, J.S., 2020. The SARS-CoV-2 Spike Protein D614G Mutation Shows Increasing Dominance and May Confer a Structural Advantage to the Furin Cleavage Domain. Preprints.

Wang, R., Chen, J., Hozumi, Y., Yin, C. and Wei, G.W., 2020. Decoding asymptomatic COVID-19 infection and transmission. The journal of physical chemistry letters, 11(23), pp. 10007–10015.

Weissman, D., Alameh, M.G., de Silva, T., Collini, P., Hornsby, H., Brown, R., LaBranche, C.C., Edwards, R.J., Sutherland, L., Santra, S. and Mansouri, K., 2020. D614G spike mutation increases SARS CoV-2 susceptibility to neutralization. Cell host & microbe.

Yang, P. and Wang, X., 2020. COVID-19: a new challenge for human beings. Cellular & molecular immunology, 17(5), pp. 555–557.

Zhang, L., Jackson, C.B., Mou, H., Ojha, A., Peng, H., Quinlan, B.D., Rangarajan, E.S., Pan, A., Vanderheiden, A., Suthar, M.S. and Li, W., 2020. SARS-CoV-2 spike-protein D614G mutation increases virion spike density and infectivity. Nature communications, 11(1), pp. 1–9.

Zhang, X., Tan, Y., Ling, Y., Lu, G., Liu, F., Yi, Z., Jia, X., Wu, M., Shi, B., Xu, S. and Chen, J., 2020. Viral and host factors related to the clinical outcome of COVID-19. Nature, 583(7816), pp. 437–440.

